# Pre-Dementia Indicators and Multidomain Vulnerabilities: Insights from AD8 Screening in Older Chinese Speaking Adults

**DOI:** 10.64898/2026.04.08.26350424

**Authors:** Wenpeng You, Fung Kuen Koo, Yu (Carrie) Cheng, Jiayi (Winnie) Huang, Hui (Fiona) Huang, Molly Li, Jacob Sevastidis, Hui-Chen (Rita) Chang

## Abstract

**Background:** Early recognition of dementia-related changes is critical for timely intervention. The AD8 Dementia Screening Interview (AD8) detects subtle cognitive and functional changes, yet its broader associations with health and wellbeing among Chinese-speaking older adults remain underexplored.

**Methods:** A cross-sectional study was conducted with 144 community-dwelling Chinese older adults (mean age 73.1 years; 81.3% female). Participants completed sociodemographic, health, functional, and psychosocial measures, including the AD8 and the Geriatric Depression Scale (GDS-15). Exploratory Factor Analysis (EFA) assessed the dimensionality of the AD8, and binary logistic regression examined associations between AD8 items and demographic, health, functional, and psychosocial outcomes.

**Results:** Chronic disease was prevalent (68.1 percent), and 13.2 percent reported a mental health disorder. EFA identified three domains: memory impairment, executive and interest decline, and functional recall difficulties, explaining 61.7 percent of the variance. Logistic regression showed predictive roles for judgment problems (AD8_1), repetition (AD8_3), financial difficulties (AD8_6), tool-use difficulties (AD8_4), and daily memory problems (AD8_8). Financial and executive difficulties were associated with age and mobility challenges, while repetition predicted psychological disorders and hopelessness. Judgment problems were linked to lower life satisfaction and happiness but greater helplessness. Worthlessness was predicted by financial, tool-use, and memory difficulties, whereas intact temporal recall (AD8_5) was protective. Several outcomes including boredom, low energy, and staying home were not significant.

**Conclusion:** Distinct AD8 items predicted vulnerabilities across physical, psychological, and social domains. Findings highlight the multidimensional value of the AD8 as a culturally relevant screening and risk stratification tool for community-based assessments of Chinese older adults.

**Summary Statement Implications for Practice:** *What does this research add to existing knowledge in gerontology?:* This study shows that specific AD8 items identify early multidimensional vulnerability among community-dwelling Chinese-speaking older adults. Difficulties with judgment, repetition, financial management, tool use, and daily memory were associated with functional limitations and psychosocial distress, extending the AD8 beyond dementia screening alone. The identification of three AD8 domains supports a broader understanding of early cognitive change as involving cognitive, functional, and emotional processes. The findings contribute culturally specific evidence from an under-researched population in gerontological research.

*What are the implications of this new knowledge for nursing care with older people?:* For nursing practice, the AD8 provides a brief, feasible tool to support holistic assessment in community and aged care settings. Key AD8 indicators can guide nurses in identifying older people at risk of functional decline and emotional vulnerability, enabling earlier, person-centred interventions. The findings highlight the importance of culturally and linguistically appropriate assessment when caring for diverse ageing populations.

*How could the findings be used to influence policy or practice or research or education?:* The results support integrating brief cognitive screening into routine nursing assessments and community-based aged care services to promote early identification and ageing in place. Nursing education should emphasise interpreting cognitive screening within psychosocial and cultural contexts. Longitudinal research is needed to assess intervention effectiveness.

**Key Points:**

➢ Early cognitive changes matter for older Chinese-speaking adults, because difficulties with judgment, repetition, financial management, and tool use (AD8 domains) were consistently linked to poorer functional and psychosocial outcomes.
➢ Beyond dementia screening, the AD8 proved useful for detecting vulnerabilities in wellbeing and daily functioning, extending its role beyond diagnostic sensitivity.
➢ A cultural focus is vital, as this study is among the first to examine AD8 associations in older Chinese-speaking adults, underscoring the need for culturally tailored screening.
➢ The psychosocial impact of cognitive changes was evident, with strong associations to helplessness, worthlessness, and reduced life satisfaction, reinforcing the overlap between cognitive and emotional health.
➢ In practice, integrating AD8 screening into community and primary care could help identify at-risk individuals early and support targeted interventions in culturally and linguistically diverse populations.

## Introduction

Dementia is a growing global health challenge, affecting more than 55 million people worldwide, with prevalence expected to rise substantially as populations age (WHO 2021). Early identification of cognitive decline is essential for timely diagnosis, care planning, and prevention of avoidable functional deterioration (He, Dieciuc et al. 2023). Early dementia-related changes often manifest subtly through behavioural, functional, or psychosocial shifts rather than overt memory loss, underscoring the need for screening tools that capture early and multidimensional indicators of decline (Bradford, Upchurch et al. 2011, Vik, Kociński et al. 2023, Alzola, Carnero et al. 2024).

The AD8 Dementia Screening Interview (AD8) is a brief informant- or self-report instrument designed to detect perceived changes across eight domains, including memory, judgment, interest, executive functioning, and daily activities (Galvin, Roe et al. 2005, Galvin, Roe et al. 2006). It has demonstrated good ability to distinguish normal ageing from early dementia (Yang, Galvin et al. 2011, Razavi, Tolea et al. 2014). However, existing research has largely focused on diagnostic accuracy, with limited attention to how individual AD8 items relate to broader health, functional, and psychosocial outcomes. Examining these item-level associations may extend the utility of the AD8 beyond screening to the identification of multidimensional vulnerability in later life (Galvin, Roe et al. 2005, Milne, Culverwell et al. 2008). In this context, the AD8 captures early changes closely aligned with intrinsic capacity, encompassing cognitive, emotional, and functional abilities that are central to maintaining independence and healthy ageing.

Older Chinese-speaking populations represent an important yet under-researched group. Barriers to dementia care, including language-related health inequities, limited culturally tailored services, and mental health stigma, may delay recognition and support (Wu, Lombardo et al. 2010, Kenning, Daker-White et al. 2017). In addition, reliance on Western-developed cognitive tools may underestimate impairment in linguistically diverse populations, highlighting the importance of culturally appropriate screening approaches(Chin, Negash et al. 2011, O’Driscoll and Shaikh 2017). Given the growing Chinese diaspora in countries such as Australia, Canada, and the United States, extending dementia screening research in this context is increasingly important (Dong, Chang et al. 2010, Elliott, Di Minno et al. 2014).

Psychosocial wellbeing, including emotional vulnerability, perceived worth, and life satisfaction, is closely linked to cognitive and functional change in later life (Jongenelis, Pot et al. 2004, Luppa, Luck et al. 2010, Henry, Coundouris et al. 2023). While tools such as the Geriatric Depression Scale capture these dimensions (Yesavage, Brink et al. 1982, Mitchell, Bird et al. 2010), little is known about how specific AD8-reported difficulties, such as impaired judgment, repetitive questioning, or reduced interest in activities, are associated with emotional vulnerability or perceptions of worthlessness (Galvin, Roe et al. 2006, Milne, Culverwell et al. 2008).

Functional limitations, including fatigue, mobility difficulties, and illness burden, frequently accompany early cognitive impairment. Difficulties managing complex daily tasks, such as financial affairs or tool use, often precede loss of independence (Avlund, Rantanen et al. 2006, Pérès, Helmer et al. 2008). However, the extent to which AD8 items reflect physical and functional vulnerability in Chinese-speaking older adults remains unclear.

This study examined associations between individual AD8 items and demographic, health, functional, and psychosocial outcomes in community-dwelling Chinese-speaking older adults. Using a cross-sectional design, exploratory factor analysis assessed AD8 dimensionality, and logistic regression examined predictors of outcomes including chronic disease, mobility limitations, life satisfaction and psychosocial vulnerability indicators. We hypothesised that AD8 items would differentially predict outcomes across domains, extending the relevance of the AD8 beyond dementia screening to multidimensional risk identification in older Chinese-speaking populations.

## 2. Methods

### 2.1 Study design and participants

This cross-sectional study was conducted in community settings in Australia where Chinese-speaking older adults engage in cultural, social, and recreational activities. Recruitment was facilitated through Chinese community organisations, cultural associations, and informal networks to ensure culturally appropriate access.

Purposive sampling was used to recruit community-dwelling adults aged ≥60 years who self-identified as Chinese. Community leaders supported recruitment through posters, word-of-mouth, and group meetings. Eligibility required adequate Mandarin literacy or verbal comprehension to complete the questionnaire independently or with minimal assistance. Individuals with severe cognitive impairment or acute illness were excluded. Of 150 individuals recruited, 144 provided valid responses (response rate 96.0%).

### 2.2 Sample size

Sample size adequacy was determined for both exploratory factor analysis (EFA) and logistic regression. The achieved sample of 144 participants exceeded recommended participant-to-item ratios for EFA of the eight AD8 items and provided sufficient events to support multivariable logistic regression analyses across most outcomes.

### 2.3 Measures

Data were collected using a structured bilingual (English–Chinese) self-administered questionnaire designed for cultural and linguistic relevance. Sociodemographic variables included age, gender, education, language, and country of birth. Health measures captured chronic conditions, diagnosed mental health disorders, and unintentional weight loss.

Cognitive changes were assessed using the eight-item AD8 Dementia Screening Interview, adapted for self-report to capture perceived changes in judgment, memory, repetition, temporal orientation, financial management, and daily functioning. The AD8 was administered as a self-report measure to enhance feasibility in community-based settings and linguistic accessibility among Chinese-speaking older adults.

Psychosocial wellbeing indicators, including life satisfaction, emotional vulnerability, and perceived worth, were assessed using selected items derived from the 15-item Geriatric Depression Scale. Individual items were examined as indicators of psychosocial vulnerability rather than as a diagnostic measure of depression, consistent with the study’s focus on multidimensional functional risk.

Functional and health outcomes included fatigue, mobility limitations, doctor-diagnosed illness, and clinically significant weight loss (greater than five percent in the past 12 months). The questionnaire was piloted for clarity and cultural appropriateness prior to data collection.

### 2.4 Data collection

Data were collected between October and December 2024 in collaboration with Chinese community organisations. Participants received written study information and provided informed consent prior to participation, with bilingual research assistants available to assist where required. Questionnaires were completed in small-group sessions or scheduled appointments, taking approximately 20–30 minutes. Responses were checked for completeness, de-identified, and entered into SPSS with double-entry verification.

### 2.5 Data analysis

Data were analysed using IBM SPSS Statistics (Version 31). Descriptive statistics summarised participant characteristics and outcome measures. The dimensional structure of the AD8 was examined using exploratory factor analysis with principal axis factoring and oblimin rotation, and internal consistency was assessed using Cronbach’s alpha.

Associations between individual AD8 items and demographic, health, functional, and psychosocial outcomes were examined using binary logistic regression. All AD8 items were entered simultaneously using the enter method. Model fit was assessed using omnibus tests and Nagelkerke’s R^2^. Results are reported as odds ratios (ORs) with 95% confidence intervals (CIs). Statistical significance was set at p < .05, with p = .05–.10 considered borderline.

### 2.6 Ethics

The study was approved by the Human Research Ethics Committee at the xxxxx (Approval No. xxxx) and conducted in accordance with the Declaration of Helsinki. All participants provided written informed consent and were informed of their right to withdraw at any time.

### 2.7 Use of AI-assisted tools

AI-assisted language tools were used solely to support clarity, grammar, and academic expression during manuscript preparation. No AI tools were used for study design, data collection, data analysis, result generation, or interpretation, and all scientific content, analyses, and conclusions were developed and verified by the authors.

## 3 Results

Among the 144 participants (mean age 73.1 years), the sample was predominantly female and generally well educated, with approximately one-third holding a bachelor’s degree or higher. Most participants identified Chinese as their first language, spoke Chinese at home, and were born in Mainland China.

Chronic conditions were common, affecting over two-thirds of participants, with nearly one-third reporting multimorbidity. Hypertension and arthritis were the most frequently reported conditions. Diagnosed psychological or mental disorders were reported by a minority of participants.

Overall, the sample represented community-dwelling, Chinese-speaking older adults with a high burden of chronic disease and relatively low prevalence of diagnosed mental health conditions (Table 1).

**Table 1.**
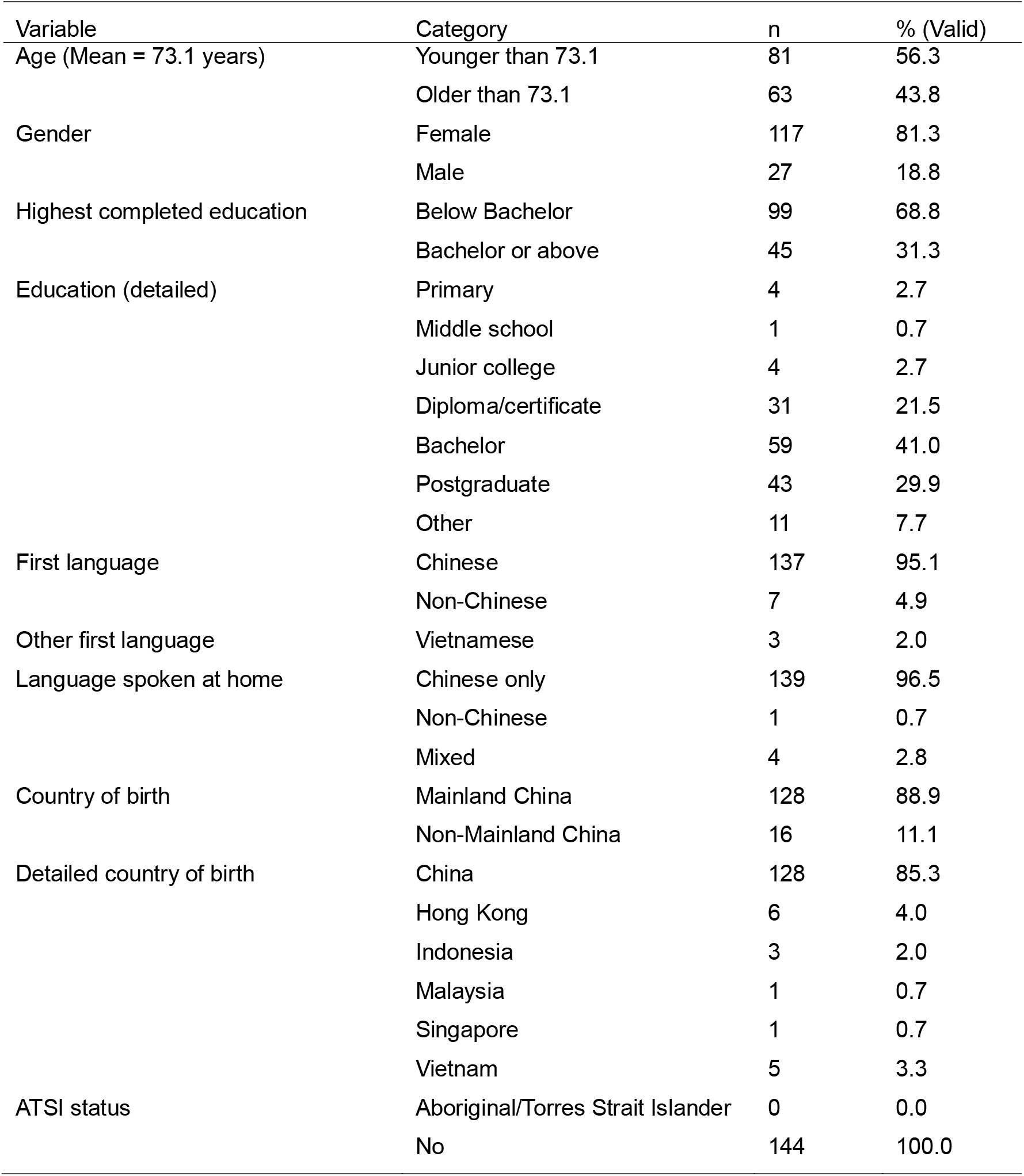
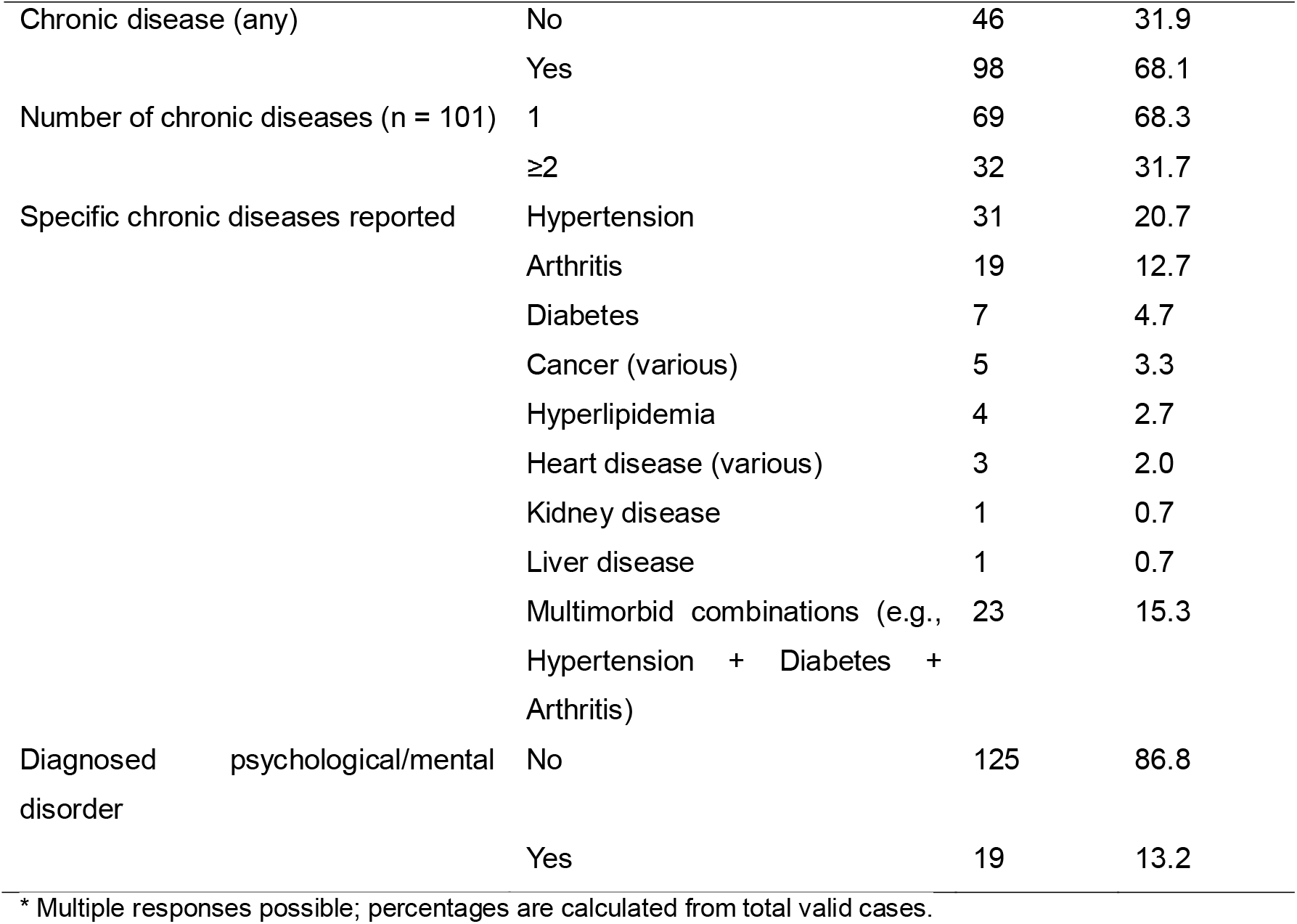
Sociodemographic and Health Characteristics of Participants (Valid n = 144)

Across the eight AD8 items, most participants reported no perceived change; however, a substantial minority endorsed changes suggestive of early cognitive or functional decline (Table 2). Difficulties related to temporal orientation, tool or appliance use, reduced interest in activities, and daily memory or thinking problems were the most frequently reported changes, each affecting approximately one-quarter to one-third of participants. Reports of judgment difficulties, repetition, financial management problems, and difficulty remembering appointments were less common but still evident in around one-fifth to one-quarter of the sample. “Don’t know” responses were infrequent across all items, indicating good item comprehensibility and response completeness.

**Table 2:** Distribution of Responses to AD8 Dementia Screening Items (N = 144)

| Variable | Description | N (Valid) | 0 = No change | 1 = Yes, a change | 2 = N/A / Don't know |
| --- | --- | --- | --- | --- | --- |
| AD8_1 | Problems with judgment (e.g., making decisions, bad financial decisions, thinking issues) | 144 | 109 (75.7%) | 32 (22.2%) | 3 (2.1%) |
| AD8_2 | Less interest in hobbies/activities | 144 | 99 (68.8%) | 42 (29.2%) | 3 (2.1%) |
| AD8_3 | Repeats same things (questions, stories, statements) | 144 | 104 (72.2%) | 38 (26.4%) | 2 (1.4%) |
| AD8_4 | Trouble learning how to use tools/appliances/gadgets | 144 | 97 (67.4%) | 47 (32.6%) | 0 (0.0%) |
| AD8_5 | Forgets correct month or year | 144 | 91 (63.2%) | 51 (35.4%) | 2 (1.4%) |
| AD8_6 | Trouble handling complicated financial affairs | 144 | 106 (73.6%) | 28 (19.4%) | 10 (6.9%) |
| AD8_7 | Trouble remembering appointments | 144 | 110 (76.4%) | 33 (22.9%) | 1 (0.7%) |
| AD8_8 | Daily problems with thinking and/or memory | 144 | 97 (67.4%) | 42 (29.2%) | 5 (3.5%) |

Fatigue and mobility limitations were common in the sample, whereas diagnosed illness and clinically significant weight loss were relatively uncommon (Table 3). A substantial proportion of participants reported frequent tiredness, and many experienced difficulty with stair climbing or walking longer distances, indicating functional vulnerability. In contrast, few participants reported a doctor-diagnosed illness or unintentional weight loss exceeding 5%.

**Table 3.** Distribution of Responses to Functional Status and Health Items (N = 144)

| Variable Description | N (Valid) | Categories / Responses | Frequency (%) | Missing |
| --- | --- | --- | --- | --- |
| How much of the time during the past 4 weeks did you feel tired? | 144 | 1 = Very little | 7 (4.9%) | 6 (4.0%) |
|  |  | 2 = Some of the time | 15 (10.4%) |  |
|  |  | 3 = A good bit of the time | 65 (45.1%) |  |
|  |  | 4 = Most of the time | 43 (29.9%) |  |
|  |  | 5 = All of the time | 14 (9.7%) |  |
| Difficulty walking up 10 steps without resting (unaided) | 144 | 0 = No | 84 (58.3%) | 6 (4.0%) |
|  |  | 1 = Yes | 60 (41.7%) |  |
| Difficulty walking several hundred yards (unaided) | 144 | 0 = No | 114 (79.2%) | 6 (4.0%) |
|  |  | 1 = Yes | 30 (20.8%) |  |
| Doctor ever diagnosed ≥1 listed illness | 144 | 0 = No | 134 (93.1%) | 6 (4.0%) |
|  |  | 1 = Yes | 10 (6.9%) |  |
| Percentage of weight loss | 144 | 0 = Less than 5% | 132 (91.7%) | 6 (4.0%) |
|  |  | 1 = Over 5% | 12 (8.3%) |  |

Overall, participants reported high levels of life satisfaction and positive affect; however, a substantial minority endorsed depressive symptoms and memory concerns (Table 4). While most respondents indicated feeling satisfied with life, happy, energetic, and positive about being alive, approximately one-quarter reported feelings of helplessness, boredom, or emptiness. Subjective memory concerns were common, and smaller proportions reported worthlessness, hopelessness, or social comparison concerns, indicating the presence of psychosocial vulnerability despite generally positive self-reported wellbeing.

**Table 4.** Distribution of Responses to GDS-15 Items (N = 144)

| Variable | Item Description | No n (%) | Yes n (%) |
| --- | --- | --- | --- |
| GDS_1 | Are you basically satisfied with your life? | 13 (9.0) | 131 (91.0) |
| GDS_2 | Have you dropped many of your activities or interests? | 105 (72.9) | 39 (27.1) |
| GDS_3 | Do you feel that your life is empty? | 110 (76.4) | 34 (23.6) |
| GDS_4 | Do you often get bored? | 103 (71.5) | 41 (28.5) |
| GDS_5 | Are you in good spirits most of the time? | 35 (24.3) | 109 (75.7) |
| GDS_6 | Are you afraid that something bad is going to happen to you? | 92 (63.9) | 52 (36.1) |
| GDS_7 | Do you feel happy most of the time? | 17 (11.8) | 127 (88.2) |
| GDS_8 | Do you often feel helpless? | 108 (75.0) | 36 (25.0) |
| GDS_9 | Do you prefer to stay at home rather than going out and doing new things? | 110 (76.4) | 34 (23.6) |
| GDS_10 | Do you feel that you have more problems with memory than most? | 82 (56.9) | 62 (43.1) |
| GDS_11 | Do you think it is wonderful to be alive now? | 8 (5.6) | 136 (94.4) |
| GDS_12 | Do you feel pretty worthless the way you are now? | 117 (81.3) | 27 (18.8) |
| GDS_13 | Do you feel full of energy? | 32 (22.2) | 112 (77.8) |
| GDS_14 | Do you feel that your situation is hopeless? | 134 (93.1) | 10 (6.9) |
| GDS_15 | Do you think that most people are better off than you are? | 116 (80.6) | 28 (19.4) |

Exploratory factor analysis of the eight AD8 items supported a three-factor structure, explaining 61.7% of the total variance (Table 5). Sampling adequacy was acceptable, and internal consistency of the scale was satisfactory (Cronbach’s α = .685). The factors reflected memory impairment (e.g., temporal disorientation and repetition), executive and interest decline (e.g., reduced interest, difficulty using tools, and financial management), and functional recall difficulties (e.g., remembering appointments and daily memory problems). Inter-factor correlations indicated related but distinct domains of cognitive and functional change.

**Table 5.** Factor Loadings and Communalities for AD8 Screening Items (N = 144)

| Item # | Description | Factor 1:<br>Memory<br>Impairment | Factor 2:<br>Executive &<br>Interest<br>Decline | Factor 3:<br>Functional<br>Recall<br>Difficulties | Communality<br>(h<sup>2</sup>) |
| --- | --- | --- | --- | --- | --- |
| AD8_1 | Problems with judgment (e.g., decisions, financial issues, thinking) | – | .367 | – | .159 |
| AD8_2 | Less interest in hobbies/activities | – | .770 | – | .577 |
| AD8_3 | Repeats the same things (questions, stories, statements) | .441 | – | – | .378 |
| AD8_4 | Trouble learning to use tools/appliances | – | .524 | – | .347 |
| AD8_5 | Forgets correct month or year | .836 | – | – | .712 |
| AD8_6 | Trouble handling complicated financial affairs | – | .495 | – | .247 |
| AD8_7 | Trouble remembering appointments | – | – | .690 | .459 |
| AD8_8 | Daily problems with thinking and/or memory | – | – | .588 | .457 |

Notes: Extraction method = Principal Axis Factoring; Rotation method = Oblimin with Kaiser normalization. Factor loadings below .30 are suppressed. Cronbach's $\alpha$ for the 8-item scale = .685.

Binary logistic regression analyses identified several domain-specific associations between AD8 items and demographic, health, functional, and psychosocial outcomes (Table 6). Demographic associations were limited, with older age related to financial management difficulties and gender differences observed in tool or appliance use; education, first language, and country of birth were not significant predictors.

**Table 6.** ealth, and Psychosocial Outcomes.

| erke R<sup>2</sup> | n | Significant AD8 Predictors (OR, p) |
| --- | --- | --- |
|  | 144 | AD8_6 Finances (1.98, .032) |
|  | 144 | AD8_4 Tools/appliances (0.16, .026) |
|  | 144 | None |
|  | 144 | None |
|  | 144 | None (trend: AD8_3 Repeats, .079) |
|  | 144 | None |
|  | 101 | AD8_1 Judgment (0.19, .012); AD8_3 Repeats (3.60, .029) |
|  | 144 | AD8_3 Repeats (4.10, .026) |
|  | 144 | None |
|  | 144 | AD8_4 Tools (3.06, .013); AD8_6 Finances (2.04, .031) |
|  | 144 | Borderline: AD8_2 Less interest (2.50, .059); AD8_3 Repeats (2.65, .056) |
|  | 144 | AD8_2 Less interest (9.19, .014) |
|  | 144 | None |
|  | 144 | AD8_1 Judgment (0.19, .006); borderline AD8_4 Tools (5.51, .061) |
|  | 144 | AD8_3 Repeats (3.93, .007); AD8_6 Finances (2.17, .024); borderline AD8_2 Less interest (2.40, .061), AD8_1 J |
|  | 144 | AD8_8 Daily memory (2.45, .036); borderline AD8_1 Judgment (2.19, .064) |
|  | 144 | None |
|  | 144 | AD8_1 Judgment (0.45, .049) |
|  | 144 | None (borderline AD8_5 Forgets month/year: 1.96, .104) |
|  | 144 | AD8_1 Judgment (0.19, .002); AD8_6 Finances (0.32, .018); AD8_8 Daily memory (10.92, .009) |
|  | 144 | AD8_1 Judgment (4.67, <.001) |
|  | 144 | None |
|  | 144 | AD8_6 Finances (2.80, .004); AD8_7 Appointments (4.96, .003) |
|  | 144 | None |
|  | 144 | AD8_4 Tools (8.87, .001); AD8_6 Finances (3.92, <.001); AD8_8 Daily memory (4.46, .010); AD8_5 Month/year |
|  | 144 | None |
|  | 144 | AD8_3 Repeats (5.30, .047) |
|  | 144 | AD8_6 Finances (2.25, .024); borderline AD8_3 Repeats (p = .057) |

Health and functional outcomes showed clearer patterns. Repetition and judgment difficulties were associated with multimorbidity, while repetition was also the strongest predictor of diagnosed psychological disorders. Mobility limitations were consistently associated with difficulties in tool use and financial management, and reduced interest in activities strongly predicted doctor-diagnosed illness. In contrast, weight loss outcomes were not significantly associated with AD8 items.

Psychosocial outcomes were most sensitive to AD8 indicators. Judgment difficulties were associated with lower life satisfaction, reduced positive affect, and greater helplessness. Financial and tool-use difficulties predicted poorer wellbeing and increased feelings of worthlessness, while repetition was most strongly associated with hopelessness. Daily memory problems were linked to feelings of emptiness and worthlessness, whereas intact temporal orientation showed protective effects in selected models. Several outcomes, including boredom, social withdrawal, energy, and positive appraisal of life, were not significantly predicted.

Overall, difficulties related to judgment, repetition, financial management, tool use, and daily memory emerged as the most consistent predictors across health, functional, and psychosocial domains, highlighting their relevance as early markers of multidimensional vulnerability.

## 4. Discussion

This study examined associations between cognitive and functional changes captured by the AD8 and a broad range of demographic, health, and psychosocial outcomes in older Chinese adults. While not all models were significant, several consistent and domain-specific patterns emerged, highlighting the relevance of selected AD8 items as indicators of multidimensional vulnerability.

Judgment difficulties (AD8_1) emerged as one of the most robust predictors, showing strong associations with reduced life satisfaction, increased helplessness, and lower positive affect. These findings align with previous evidence linking impaired executive functioning to reduced psychological resilience, emotional regulation, and social participation in older adults experiencing cognitive decline (Bradford, Upchurch et al. 2011, WHO 2021, He, Dieciuc et al. 2023, Vik, Kociński et al. 2023, Alzola, Carnero et al. 2024). The observed associations suggest that judgment impairment may reflect combined cognitive and affective vulnerability rather than isolated cognitive dysfunction (Galvin, Roe et al. 2005, He, Dieciuc et al. 2023).

Repetitive speech and behaviour (AD8_3) were strongly associated with psychological disorders and hopelessness, reinforcing their role as salient behavioural markers of vulnerability. Although the relationship between depression and dementia risk is well established (Yang, Galvin et al. 2011), repetition itself has received less attention as a specific indicator of emotional distress. Existing literature suggests that repetitive behaviours in dementia may reflect agitation, anxiety, or environmental stressors, and are associated with caregiver burden and emotional instability (Milne, Culverwell et al. 2008, Wu, Lombardo et al. 2010, Razavi, Tolea et al. 2014, Kenning, Daker-White et al. 2017). Our findings indicate that repetition may warrant greater consideration as an early behavioural signal of psychosocial risk, although causal pathways require further investigation (Chin, Negash et al. 2011).

Financial management difficulties (AD8_6) were among the most consistent predictors across functional and psychosocial outcomes, including mobility limitations, memory concerns, and feelings of worthlessness. Financial incapacity is widely recognised as one of the earliest functional impairments in dementia and is closely linked to autonomy and self-esteem (Elliott, Di Minno et al. 2014, O’Driscoll and Shaikh 2017). Importantly, executive dysfunction related to late-life depression may also impair financial capacity in the absence of dementia (Dong, Chang et al. 2010, Henry, Coundouris et al. 2023), underscoring the need for careful interpretation. As financial skills deteriorate progressively across dementia stages (Jongenelis, Pot et al. 2004), future refinement of AD8 financial items to capture specific financial behaviours may enhance its ability to distinguish dementia severity rather than presence alone.

Tool-use difficulties (AD8_4) were associated with mobility limitations, gender differences, and psychosocial vulnerability. Instrumental activities of daily living, including tool and appliance use, are highly sensitive to early cognitive decline and may reflect both cultural role expectations and functional deterioration (Yesavage, Brink et al. 1982, Luppa, Luck et al. 2010). These findings reinforce the importance of IADL-focused assessment in early cognitive screening.

Daily memory and thinking problems (AD8_8) were associated with feelings of emptiness and worthlessness, consistent with evidence linking subjective memory complaints to reduced quality of life and emotional vulnerability (Royall, Palmer et al. 2004, Mitchell, Bird et al. 2010). In this study, individual items derived from the Geriatric Depression Scale were used to capture psychosocial vulnerability rather than to indicate depressive disorder, in line with a multidimensional functional risk framework. Paradoxical associations with positive affect observed in some models may reflect adaptive coping, reporting biases, or culturally shaped interpretations of memory change (Avlund, Rantanen et al. 2006, Pérès, Helmer et al. 2008). In some Chinese cultural contexts, memory decline is often viewed as a normal aspect of ageing rather than pathology (Lee, Choi et al. 2020, Wei, Huang et al. 2020, Lee, Avila-Rieger et al. 2023), which may attenuate emotional distress during early symptom recognition (Shariati, Nasiri et al. 2024).

Not all outcomes were significantly predicted by AD8 items. Measures such as boredom, social withdrawal, and energy demonstrated weaker or unstable associations, suggesting that some GDS items may be less sensitive to cognitive predictors or influenced by cultural response patterns (Rajtar-Zembaty, Sałakowski et al. 2017, Frau, Jonaitis et al. 2025). Nonetheless, these psychosocial factors remain clinically relevant given their established links with geriatric depression, a recognised mediator of dementia risk (Ownby, Crocco et al. 2006, Shariati, Nasiri et al. 2024).

Overall, this study demonstrates that specific AD8 domains, particularly judgment, repetition, financial management, tool use, and daily memory, extend beyond cognitive screening to predict psychosocial vulnerability and functional burden. By mapping these domains to broader health and wellbeing outcomes in a Chinese immigrant cohort, our findings extend existing AD8 validation work (Mitchell, Bird et al. 2010, Hamdy, Lewis et al. 2018) and support the need for culturally sensitive screening approaches that integrate cognitive, functional, and emotional dimensions (Ryan, Tainsh et al. 1988, Tetsuka 2021). These insights may inform more targeted, community-based interventions to support healthy ageing in culturally and linguistically diverse populations (Springate and Tremont 2014).

### Implications for Cognitive Health and Ageing in Place

The findings have important implications for cognitive health promotion and ageing in place, consistent with the WHO Healthy Ageing framework (Dotson, Beydoun et al. 2010). Associations between key AD8 domains, particularly judgment, repetition, financial management, and tool use, and poorer psychosocial and functional outcomes underscore the need to safeguard intrinsic capacity across cognitive, emotional, and functional domains. Integrating brief cognitive screening tools such as the AD8 into routine primary and community care may facilitate earlier identification of vulnerability and support timely, targeted interventions. Community nurses, general practitioners, and aged care assessors are well placed to embed such screening within routine assessments, care planning, and referral pathways, particularly for culturally and linguistically diverse populations.

Equally important is the development of enabling environments through culturally and linguistically appropriate approaches (Marson, Sawrie et al. 2000, Dotson, Beydoun et al. 2010). Given the predominantly Chinese-speaking sample, health promotion strategies should be culturally congruent and inclusive of family and community networks to enhance engagement, reduce stigma, and support sustained participation in care (Mackin and Areán 2009, Tang 2021). The links identified between cognitive change, mobility, psychological wellbeing, and life satisfaction also reinforce the central role of functional ability, shaped by the interaction between health and environment, in determining ageing in place outcomes. Embedding cognitive monitoring within broader community health initiatives and home care packages may therefore help sustain independence, delay institutionalisation, and improve quality of life among culturally diverse ageing populations (Morin, Gonzales et al. 2019).

## Conclusion

This study demonstrates that specific cognitive and functional changes identified by the AD8 are associated with demographic, health, functional, and psychosocial outcomes in older Chinese-speaking adults. Difficulties with judgment, repetition, financial management, and tool use consistently predicted poorer wellbeing, greater psychological distress, and functional limitations, while daily memory problems were linked to feelings of emptiness and worthlessness. These findings extend the utility of the AD8 beyond dementia screening to the identification of early multidimensional vulnerabilities that may compromise healthy ageing.

The results also underscore the importance of culturally sensitive approaches to cognitive assessment in immigrant populations, where language, cultural context, and health inequities may influence symptom recognition and care pathways. Although causal inferences are limited by the cross-sectional design, the observed associations suggest that early identification of key AD8 domains may support timely, targeted interventions. Integrating cognitive, functional, and psychosocial screening within community and primary care settings may enhance resilience and wellbeing in culturally and linguistically diverse ageing populations.

### Strengths and Limitations

A key strength of this study is its focus on older Chinese-speaking adults, a population underrepresented in dementia and ageing research despite its growing presence in Australia and other Western countries. The use of culturally adapted instruments, including the AD8 and psychosocial wellbeing items derived from the GDS-15, enabled assessment of cognitive, functional, and psychosocial wellbeing in a linguistically appropriate manner. Methodological strengths include an adequate sample size, the use of multivariable logistic regression across multiple outcome domains, and exploratory factor analysis to support the multidimensional structure of the AD8.

Several limitations should be acknowledged. The cross-sectional design precludes causal inference, and self-report measures may be subject to recall or reporting bias, particularly among participants with cognitive impairment. Some regression models showed instability or non-significant results, likely reflecting sparse data in certain outcome categories. In addition, recruitment through Chinese community networks may limit generalisability to other culturally and linguistically diverse groups or to older adults outside community settings.

Despite these limitations, the study provides novel evidence linking early AD8-identified cognitive and functional changes with health and psychosocial outcomes, supporting the clinical and public health relevance of the AD8 in culturally diverse ageing populations.

## Data Availability

All data produced in the present study are available upon reasonable request to the authors.

## Acknowledgments

We sincerely thank the Chinese Australian Services Society (CASS) and Arabic Council Australia (ACA) for their essential support in participant recruitment and data collection. Their staff provided invaluable assistance with survey administration and cultural liaison, enabling successful engagement with Mandarin/Cantonese and Arabic-speaking communities. This research would not have been possible without their partnership.

## Notes

**Declaration of conflict of interest:** The Authors declare that there is no conflict of interest.

**Declaration of Competing Interest** The authors declare that they have no conflicts of interest relevant to this work.

**Funding** This research was supported by a Seed Grant from the School of Nursing and Midwifery, Western Sydney University.

**Declaration of generative AI and AI-assisted technologies in the writing process** During the preparation of this manuscript, the lead author employed OpenAI’s ChatGPT (GPT-5) to support language refinement and improve clarity of expression. Following this process, all authors critically reviewed, revised, and approved the content, and take full responsibility for the accuracy, integrity, and originality of the final manuscript.

### Competing Interest Statement

The authors have declared no competing interest.

### Funding Statement

This research was supported by a Seed Grant from the School of Nursing and Midwifery, Western Sydney University.

### Author Declarations

The Human Research Ethics Committee of the University of New England granted ethics approval (Approval number: HE22-088).

